# The IeDEA Data Exchange Standard: a common data model for global HIV cohort collaboration

**DOI:** 10.1101/2020.07.22.20159921

**Authors:** Stephany N. Duda, Beverly S. Musick, Mary-Ann Davies, Annette H. Sohn, Bruno Ledergerber, Kara Wools-Kaloustian, Catherine C. McGowan, Nicola J. Maxwell, Azar Kariminia, Cam Ha Dao Ostinelli, Brenna C. Hogan, Qiuhu Shi, Karen Malateste, Ruth L. Goodall, Dennis Karsten Kristensen, Erik V. Hansen, Carolyn F. M. Williams, Judith T. Lewis, Constantin T. Yiannoutsos

## Abstract

**Objective:** To describe content domains and applications of the IeDEA Data Exchange Standard, its development history, governance structure, and relationships to other established data models, as well as to share open source, reusable, scalable, and adaptable implementation tools with the informatics community.

**Methods:** In 2012, the International Epidemiology Databases to Evaluate AIDS (IeDEA) collaboration began development of a data exchange standard, the IeDEA DES, to support collaborative global HIV epidemiology research. With the HIV Cohorts Data Exchange Protocol as a template, a global group of data managers, statisticians, clinicians, informaticians, and epidemiologists reviewed existing data schemas and clinic data procedures to develop the HIV data exchange model. The model received a substantial update in 2017, with annual updates thereafter.

**Findings:** The resulting IeDEA DES is a patient-centric common data model designed for HIV research that has been informed by established data models from US-based electronic health records, broad experience in data collection in resource-limited settings, and informatics best practices. The IeDEA DES is inherently flexible and continues to grow based on the ongoing stewardship of the IeDEA Data Harmonization Working Group with input from external collaborators. Use of the IeDEA DES has improved multiregional collaboration within and beyond IeDEA, expediting over 95 multiregional research projects using data from more than 400 HIV care and treatment sites across seven global regions. A detailed data model specification and REDCap data entry templates that implement the IeDEA DES are publicly available on GitHub.

**Conclusions:** The IeDEA common data model and related resources are powerful tools to foster collaboration and accelerate science across research networks. While currently directed towards observational HIV research and data from resource-limited settings, this model is flexible and extendable to other areas of health research.

**Highlights:**

- The IeDEA Data Exchange Standard is a data model for HIV epidemiology research.
- The model has expedited 95 projects using data from >400 HIV clinics worldwide.
- A browsable and adaptable version and data collection templates are available online.

## Background and Significance

Shared standards for representing and exchanging data are essential in collaborative research, particularly in the area of HIV/AIDS where multi-site, multi-national, and multiregional collaborations study the global epidemic. To facilitate this work and increase the efficiency of data sharing, collaborating researchers must exchange data using common data models (CDMs), which standardize data tables and variable names, matched code lists, data definitions, and formatting guidelines. Researchers have designed generalized CDMs that capture the broad range of clinical data generated by electronic health record systems and enable rapid data aggregation and distributed analyses across sites [1–4]. Despite their flexibility, generalized data models are not suited to all types of research collaborations: they lack dedicated variables for disease-specific concepts and may not account for varying levels of precision in clinical data obtained in particular in resource-limited settings. Realizing both the advantages of standardized data sharing and the extensive challenges of collecting, harmonizing, processing, validating, and analyzing HIV-specific data obtained from diverse sources, we developed a robust common data model for HIV data exchange that addresses the local context of data collection and the needs of sites from diverse geographic and resource backgrounds.

The International Epidemiology Databases to Evaluate AIDS (IeDEA) consortium is a collaboration of seven regional HIV observational research networks in North America, Latin America and the Caribbean, the Asia-Pacific, and four regions in sub-Saharan Africa (***Figure 1***) [5]. IeDEA was launched in 2006 with support from the U.S. National Institutes of Health (NIH) to conduct global research to address high priority HIV/AIDS research questions [6]. Each of the seven IeDEA regions collates data in a variety of different formats, from participating HIV clinics in its geographic area and pools them in longitudinal databases. In most cases, these data are derived from routine patient care and include demographics, clinical visit measurements such as height and weight, HIV-related diagnoses and laboratory values, plus dosages and dates of antiretroviral and other medications. Data sources include electronic health record systems (e.g., OpenMRS, local or proprietary systems)[7,8], research databases (e.g., MS Access, SPSS, REDCap)[9–11], and paper clinic charts. Regional Data Centers are responsible for receiving data from participating HIV clinics, combining the data into a regional database for local and regional analysis, conducting data quality checks, and sending data to other IeDEA regions to be combined for multiregional research. NIH developed this federalized structure to build cultures of trust where data sharing was initiated close to the point of data creation and researchers could support constituent countries’ research needs, thereby increasing the collaboration’s flexibility and inclusiveness. This process has been refined over time, resulting in data from almost two million people living with HIV currently contained in regional databases.

**Figure 1:**
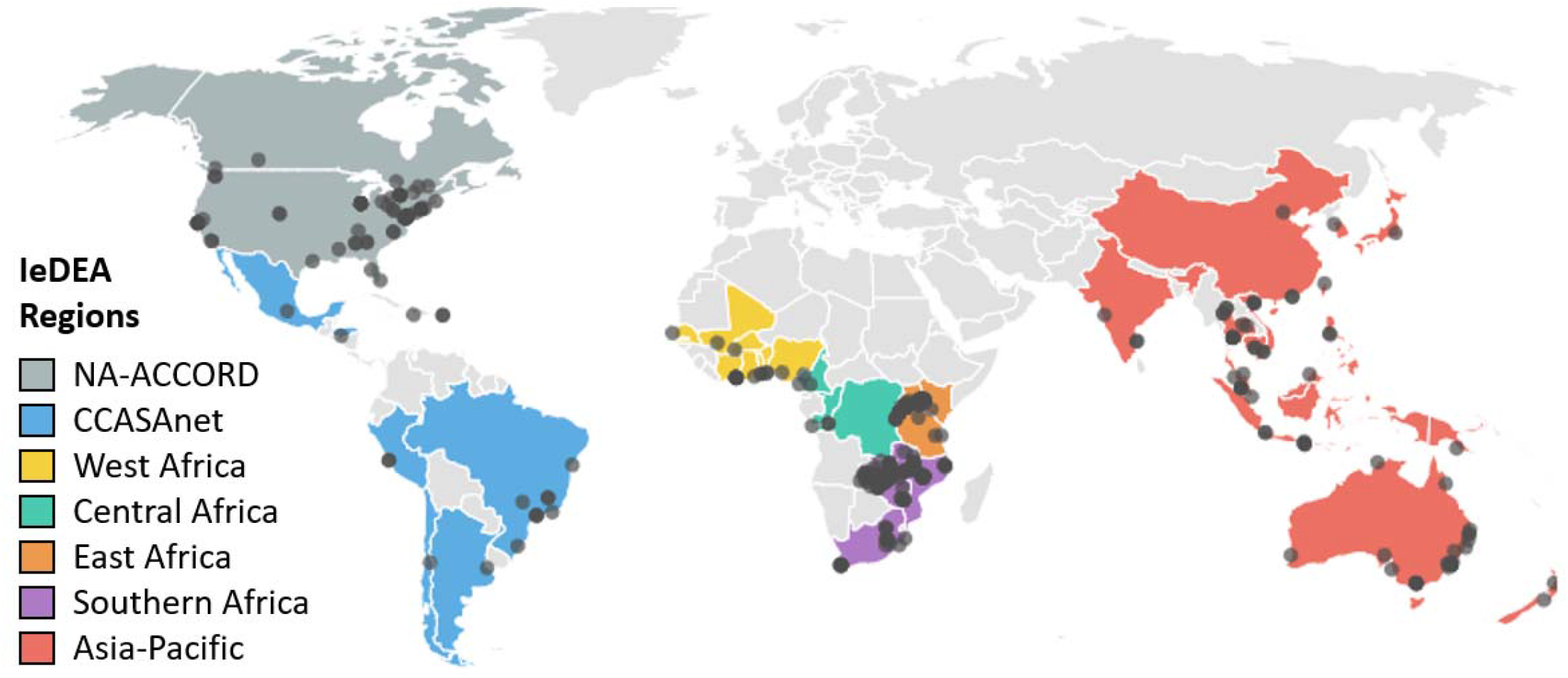
Map of the seven IeDEA regions and participating HIV clinics.

Overcoming barriers to harmonizing repositories of observational HIV data was the principal motivation behind IeDEA’s interest in common data models for HIV epidemiology. At the beginning of IeDEA, the consortium had no common data exchange standard, leading to excessive time and resources spent on reformatting, cleaning, and merging data for multiregional research. To address this challenge, our team of regional data scientists, clinicians, and analysts decided to create an easy-to-apply common data standard to facilitate HIV data exchange. Here we recount the development of the IeDEA Data Exchange Standard (IeDEA DES), outline the challenges and decisions behind its creation, and describe the evolving products of this global HIV data collaboration.

## Methods

### Development

During the July 2012 XIX International AIDS Conference in Washington, DC, IeDEA held a Data Harmonization Stakeholders’ Meeting to work towards a data standard to support global HIV epidemiology research. Attendees included over 30 clinicians, epidemiologists, informaticians, data managers, and project managers engaged in IeDEA research, along with representatives from IeDEA funding agencies. Meeting participants reviewed other available data models, including early versions of the US-based Observational Medical Outcomes Partnership (OMOP) common data model [1], the i2b2 (Informatics for Integrating Biology and the Bedside) data schema [12], and the European HIV Cohorts Data Exchange Protocol (HICDEP). HICDEP was designed in 2003 by European HIV cohorts [13] and further developed within EuroCoord, the HIV research network funded by the European Union specifically to support collaboration among European HIV cohorts [14]. At the time it was the only HIV-specific data model explicitly designed for research data sharing.

After a detailed variable-level comparison with data arising from IeDEA regions, the meeting participants advised against unconditional adoption of any existing data model. This arose from the concern that all the generalizable CDMs under consideration required complex coding and data formatting that would require substantial additional resources for data preparation, and the resulting normalized data structure was challenging for clinicians and analysts to interpret. Of all data models reviewed, HICDEP was most in line with anticipated IeDEA data management needs, but lacked sufficient flexibility in the variable definitions and coding structure. For example, at the time HICDEP did not include variables for pediatric patients or tuberculosis, and had a limited number of pregnancy-related variables. Therefore, IeDEA decided to fork the HICDEP standard by adopting core HICDEP tables and conventions while substantially expanding the variables, tables, and code lists to address broader data needs in IeDEA. A working group was assembled to define an IeDEA common data model and data exchange procedures. During a 6-month design process, IeDEA investigators consistently favored a data model that met practical, on-the-ground needs of local research teams and data managers over more abstracted data models.

The first version of the IeDEA Data Exchange Standard (IeDEA DES) was released and implemented for all IeDEA projects in 2012. However, the growing volume of multiregional collaborative work involving cross-domain HIV studies and external scientific collaborations necessitated a more complex data model. In 2015, we began an intensive effort to expand the IeDEA DES as part of the NIH-funded Big Data to Knowledge (BD2K) program. New data domains were identified through a review of IeDEA research proposals that requested data which did not exist in the previous version of the data model and through consultations with experts who helped identified data needs for emerging research priorities. All cadres of IeDEA data creators and users participated in extending the data model. Clinicians helped inform the semantics of variables, regional data managers standardized the variable definitions and syntax, and epidemiologists and statisticians provided insights into efficient data representations for analyses. Members of HICDEP and other European HIV cohorts also participated. The resulting major revision of the IeDEA DES was released in February 2017, with ongoing annual updates as of 2020.

## Results

### Content coverage

From an initial 7 tables and 45 variables in its original 2012 version, the IeDEA Data Exchange Standard has evolved to 29 tables and 269 variables in its 2020 version (***Figure 2***). The IeDEA DES covers core domains for HIV epidemiology research, including basic demographics, anthropometrics, HIV disease staging, clinic visits, medications, laboratory measurements, clinical diagnoses, hospitalizations, pregnancy data, and the results of screening and diagnostic patient questionnaires.

**Figure 2:**
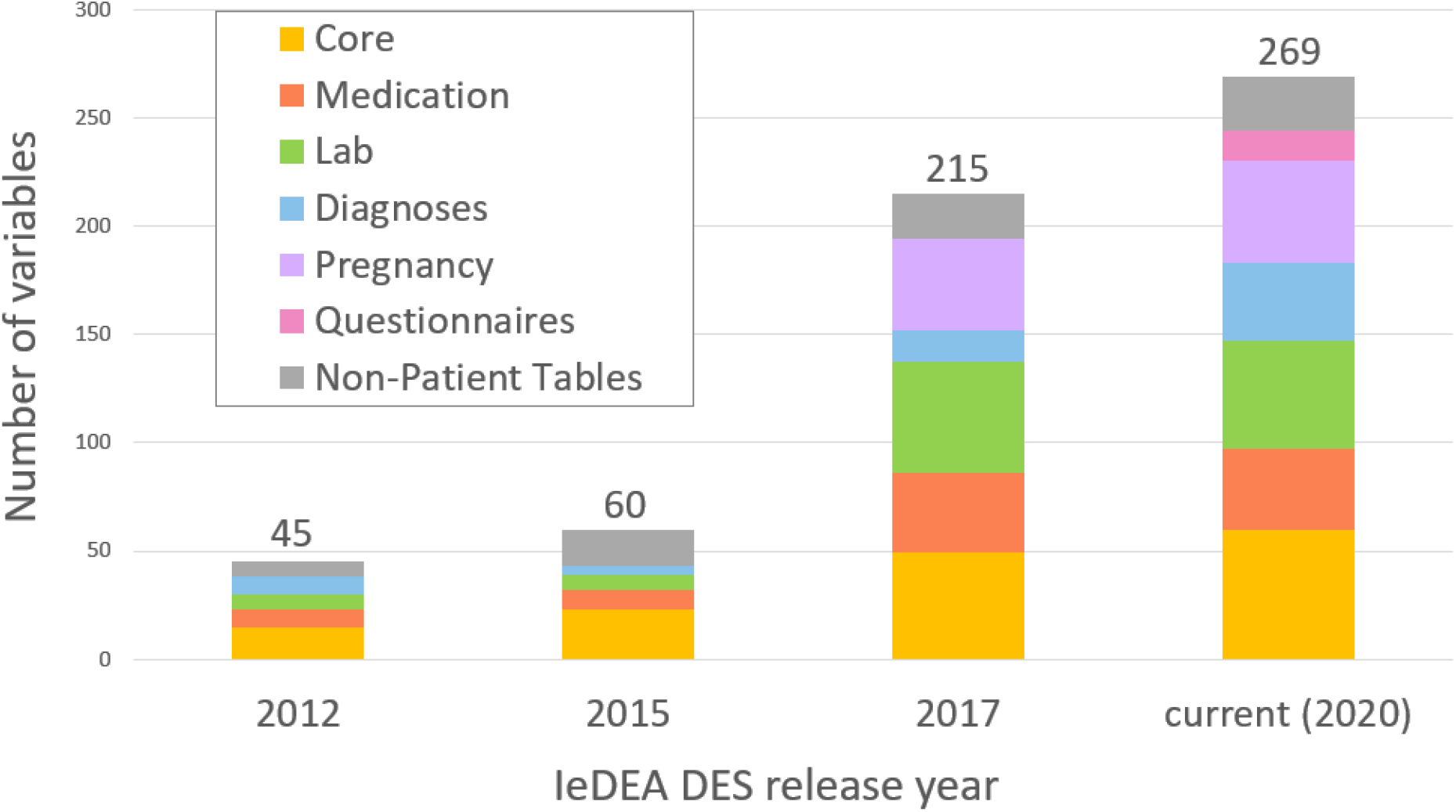
Categories and total counts of variables in the IeDEA Data Exchange Standard by major version release year

Initial patient data, including the patient identifier, birth date, sex at birth, probable route of HIV infection, and HIV clinic enrollment date, are stored in a required “basic” table (`tblBAS`), which is paired with an optional patient follow-up table that includes date of loss to follow-up from clinic, last date known alive, and date of death (`tblLTFU`). Most other patient tables, such as laboratory testing, medication, and visit tables, can contain multiple rows per patient. Through BD2K support, we added tables on pregnancy and newborn care, allowing linkage of mother-child pairs when both are enrolled in the cohort, or facilitate capture of mother-centric or child-centric data when only one member of the mother-child dyad is enrolled. Other expansions included variables for HIV status disclosure, orphan status and caregiver information for children, reasons for starting antiretroviral therapy (ART), drug resistance testing, and cancer diagnoses. ***Figure 3*** depicts a diagram of the tables and variables of the IeDEA DES data model.

**Figure 3:**
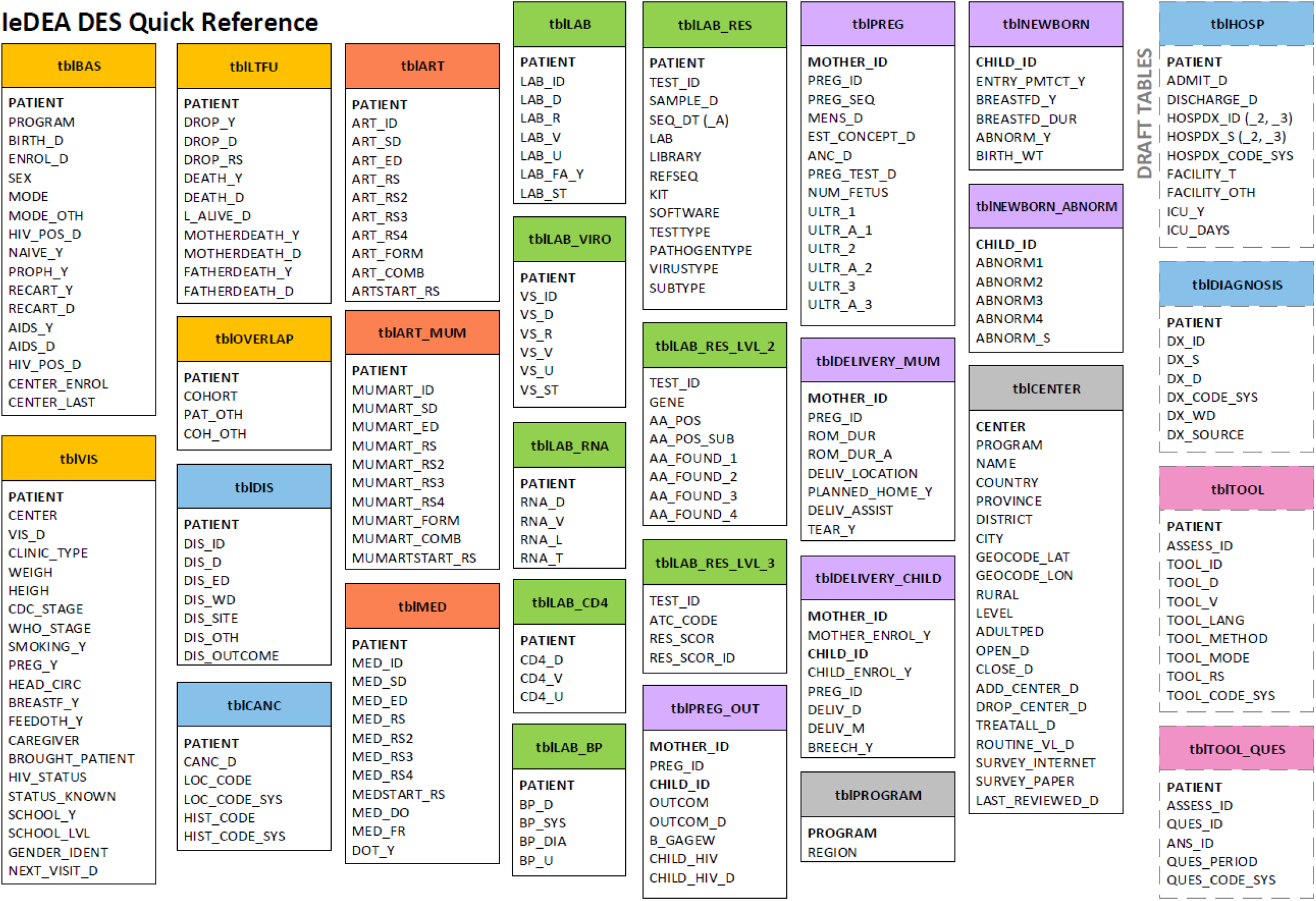
Diagram of tables and variables for the IeDEA DES data model. “Table Basic” (`tblBAS`), shown top left, is the index table containing records for all patients in the cohort. All date variables (*variablename_*D) have optional date approximation variables (*variablename_*D_A) (date approximation variables not shown). Updated versions and variable definitions are posted at iedeades.org.

Two non-patient tables, `tblCENTER` and `tblPROGRAM`, focus on data about the location of care. `tblCENTER` captures the name and location of the HIV care and treatment center, clinic population, level of care (e.g., primary to tertiary), community setting (e.g., urban or rural), and the opening and closure dates of the research dataset. `tblPROGRAM` links multiple HIV clinics that are part of the same health care program or organization.

### Design for HIV data from resource-limited settings

A key innovation of the IeDEA DES is that it supports the exchange of core HIV care data from a variety of settings ranging from a rural health clinic in South Africa, to a public hospital in Peru, to an academic medical center in Canada. The database structure is straightforward, reflects real-world clinical concepts (e.g., labs, visits, medications), and is not fully normalized, allowing it to be manipulated with limited computing resources by non-experts. Code lists include inexpensive point-of-care/rapid laboratory tests often used in resource-limited settings. Every patient-related date variable in the IeDEA DES has an optional “date approximation” field that allows data managers to specify uncertainty in the accuracy of a date. Such flexibility in defining variables, is critical for representing data extracted from clinics with less detailed medical records. Patient visits are mapped to clinics in the visits table (`tblVIS`), allowing HIV care and treatment programs to track patients who visit multiple IeDEA sites as part of routine care.

To accommodate multiple types of local data management software across IeDEA contributing sites, the IeDEA DES does not define a file format for central data transfer. DES data files can be exchanged using common formats such as CSV, generic database formats such as SQL, or proprietary data formats like SAS and Stata, depending on the needs of the data recipients and the platforms available to the sender.

### Naming conventions

IeDEA adopted many core HIV variables from HICDEP along with their naming conventions. Table names are prefixed with “tbl” to distinguish tables from variables. Variable suffixes indicate the type and format of the variable. ***Table 1*** lists the most frequently used suffixes as formalized in the IeDEA DES. These naming conventions are used to automate basic data quality checks on IeDEA datasets. For example, if the binary variable for AIDS diagnosis is coded “No” (`AIDS_Y`=0), then there should be no date in the associated `AIDS_D` field because the patient has not been diagnosed with AIDS.

**Table 1:** Variable name suffixes used in the IeDEA DES to designate data type and content

| Variable Suffix | Variable Meaning |
| --- | --- |
| **_Y** | Yes/No/Unknown |
| **_D** | Date in YYYY-MM-DD format |
| **_D_A** | Date approximation code |
| **_ID** | Identifier (except PATIENT) |
| **_OTH** | Descriptive field for “other (specify)” |
| **_S** | Descriptive field (string) for original data |
| **_R** | Result (of non-numeric test) |
| **_V** | Value (of numeric test result) |
| **_T** | Type (of facility, laboratory equipment) |
| **_U** | Unit of measurement code |
| **_RS** | Reason (for medication change, loss to follow-up, death) |

### Use of terminologies

The IeDEA DES supports the use of standardized terminologies distributed by the World Health Organization (WHO), as these coding systems have no licensing fees and are most likely to be used globally and in resource-limited settings. Medications in the antiretrovirals table (`tblART`), general medications table (`tblMED`), and drug resistance table (`tblLAB_RES_LVL_3`) are coded according to the Anatomical Therapeutic Chemical (ATC) Classification System maintained by WHO [15]. Diagnoses in the hospitalizations and diagnoses tables (`tblHOSP`, `tblDIAGNOSIS`) are coded using WHO’s International Statistical Classification of Diseases and Related Health Problems (ICD) [16]. The IeDEA Data Harmonization Working Group provides suggested subsets of ICD-10 and ATC codes to simplify coding, but other coding systems can be used if defined in a table’s coding system (`_CODE_SYS)` variables. The official lists of AIDS-defining disease diagnoses (`tblDIS`) specified by the WHO and the U.S. Centers for Disease Control and Prevention are represented using an internally managed list of 2-to-4-character strings [17,18]. This concession maintains historic compatibility with HICDEP (for CDC codes) and facilitates mapping of this critical HIV care data, as the WHO list of AIDS conditions does not map one-to-one to ICD codes. International Organization for Standardization (ISO) standards are used for date formats, country codes, and language codes [19–21].

### Governance

The IeDEA DES is maintained by the Data Harmonization Working Group, which meets monthly to assess new research proposals and their data needs. Upkeep of the DES requires approximately four hours per month, including collating new variable proposals and revising them to conform to naming conventions, researching other data models, and communicating with domain experts. During an annual cross-consortium data meeting, working group members discuss proposed IeDEA DES additions with the European HICDEP team and consider new HICDEP variables for inclusion. Backwards compatibility is preserved for all coding lists, though old variables may be deprecated with advance notice if needed to maintain consistency of the data model. A formal draft revision is prepared annually by the DHWG and circulated to all IeDEA regions for review. The revisions are presented at an IeDEA-wide meeting for approval by the IeDEA Executive Committee before release.

### Availability

The IeDEA DES is openly available for use and adaptation by other research organizations. Documentation of the IeDEA DES is provided at http://iedeades.org and recorded on fairsharing.org and the NIAID resources page [22,23]. To support use of the IeDEA DES for practical data collection, the version of the IeDEA DES described here has been implemented as a set of data collection forms for the REDCap platform. These REDCap data dictionaries, which were developed for REDCap version 8.2.0 and above, are posted on GitHub at https://github.com/IeDEA in CDISC ODM-XML format, along with implementation guidelines and a REDCap data dictionary for the communally maintained `tblCENTER`.

### Impact

Data from more than 400 HIV care and treatment clinics participating in IeDEA have been successfully mapped to the DES for multiregional data exchange. Since its launch in 2012, the DES has facilitated data exchange for more than 95 projects and 49 publications across IeDEA regions and with other external collaborators such as the World Health Organization and the Joint United Nations Programme on HIV/AIDS (UNAIDS)[24–26]. Multiregional IeDEA data also have been used by other research groups, including the Collaborative Initiative for Paediatric HIV Education and Research (CIPHER)[27–29], the Collaboration of Observational HIV Epidemiological Research Europe Study (COHERE) [30,31], and the ALPHA network [32]. IeDEA also contributes data and scientific expertise to mathematical modeling consortia such as the Cost-Effectiveness of Preventing AIDS Complications (CEPAC) modeling group [33,34] and the UNAIDS Reference Group on Estimates, Modelling and Projections [35–40]. Clear benefits of the IeDEA DES have included substantial increases in data quality observed by data managers and analysts, tangible time savings from standardization when performing repeated data processing, and increased collaboration and perceived added value among participating organizations. Three IeDEA regions have adopted the IeDEA DES as their principal data storage format and the European HICDEP collaboration has incorporated most of IeDEA’s additions into its own data model.

## Discussion

The IeDEA DES is an accessible data standard with an active community of contributors, a proven tool to improve efficiency and facilitate global HIV research collaborations, and a resource for meaningful multiregional collaboration within and beyond IeDEA. Its use in IeDEA has facilitated numerous projects, publications, and collaborations. IeDEA chose to develop a patient-centric, disease-centric data model rather than using a generalized CDM to reduce resource utilization. Mapping clinical data to a generalized CDM and extracting them for analysis often involves extensive data abstraction, transformation, and recoding steps and consequently more personnel time, data management expertise, and data infrastructure. Our approach towards common data model adoption reflects our “on the ground” experience with data challenges in global collaborations.

Plans for future growth of the IeDEA DES include developing new tables and variables to better represent patient antiretroviral treatment adherence, entry points into care (e.g., voluntary counseling and testing, antenatal clinics), and participation in research trials, as well as HIV care program characteristics. As HIV care is transformed from acute infectious disease management to chronic disease care, the IeDEA DES will expand to include more tables and variables to cover non-communicable disease outcomes. The model is also being expanded to include tuberculosis disease and care. Expansion of the IeDEA DES continues to be informed by common data models originally developed for electronic health record data (e.g., Observational Medical Outcomes Partnership (OMOP) CDM, PCORnet Distributed Research Network CDM [1–3]), as well as electronic health record database schemas designed for HIV care, such as the Academic Model Providing Access to Healthcare (AMPATH) Medical Record System schema and the HIV minimum dataset proposed by Tierney et al. [41,42].

The structure of the IeDEA DES allows for the development of research support software, and we are currently developing software platforms where IeDEA DES data can be loaded, quality-checked, and summarized. Through the continuous expansion and improvement of its data domains, and because of its proven utility in worldwide collaborations and accessibility, the IeDEA DES is positioned as a complete solution for HIV-specific data collection, definition, and storage in the “big epidemiology data” context.

## Conclusions

The IeDEA DES is an accessible data exchange standard that continues to be expanded based on feedback from global collaborators and shaped by evolving HIV epidemics and new research interests. The IeDEA data model can be leveraged and adapted by other research collaborations seeking to implement a defined structure for disease-specific, patient-centric research data exchange. The ongoing data standardization efforts promoted by IeDEA and the increased awareness of the benefits of global data harmonization are intended to meaningfully strengthen collaborations studying the impact of the global response to the HIV epidemic.

## Data Availability

The manuscript does not reference data.

https://github.com/IeDEA

## Acknowledgments

The authors would like to acknowledge the support of the members of the IeDEA Data Harmonization Working Group, the IeDEA Executive Committee, the HIV Cohorts Data Exchange Protocol collaboration, and other groups that have contributed to the development of the IeDEA DES over time. The authors also thank Dr. Pernille Iversen, Mr. Jesper Kjaer, Mr. Larry Riggen, Dr. Rosemary McKaig, and Dr. Firas Wehbe for their support in the original creation of the IeDEA Data Exchange Standard.

